# Variation in Tolvaptan Prescribing for Autosomal Dominant Polycystic Kidney Disease in the United Kingdom and Its Impact on Quality of Life and Costs

**DOI:** 10.64898/2026.04.04.26350154

**Authors:** Mathew Gittus, David Pitcher, Alicia O’Cathain, Albert CM Ong, Roslyn Simms, James Fotheringham

**Affiliations:** University of Sheffield, School of Medicine and Population Health, Sheffield, United Kingdom; National Registry of Rare Kidney Diseases, Bristol, United Kingdom; Sheffield Kidney Institute, Sheffield, United Kingdom

## Abstract

**Background and hypothesis:** Autosomal dominant polycystic kidney disease (ADPKD) affects over 12 million people worldwide including an estimated 30,000–70,000 in the United Kingdom (UK). Tolvaptan is the only disease-modifying therapy approved for rapidly progressing disease. Despite national guidance, prescribing rates were hypothesised to vary by kidney centre. Treatment may not always align with guidelines: some patients eligible for tolvaptan may not be initiated, while other patients initiated on tolvaptan may not meet eligibility criteria. This may have important consequences for healthcare costs and health-related quality of life.

**Methods:** The National Registry of Rare Kidney Diseases (RaDaR) collects longitudinal data from UK NHS kidney centres. This retrospective cohort study used routinely collected data (2016-2023) to examine tolvaptan prescribing across kidney centres. Kidney centre-level initiation patterns were described, assessed using mixed-effects logistic regression and visualised with funnel plots. Cost-effectiveness analyses combined observed prescribing practices under likely negotiated commercial discounts to estimate costs and quality-adjusted life year (QALY) consequences of prescribing at the national level.

**Results:** Our study included 3,609 people with ADPKD from 72 kidney centres. Patients eligible for tolvaptan who were not initiated accounted for 34.8% (292/839). Across centres, five (6.9%) initiated tolvaptan significantly more than expected among eligible participants, while one centre (1.4%) initiated significantly less. Nationally, this could result in up to £53.7 million in lost savings (assuming a 60% medication price reduction) and result in up to 1,245 lost QALYs. Patients initiated on tolvaptan who were not eligible accounted for 26.1% (103/395). Only one centre had significantly fewer eligible patients than expected among initiated patients. Nationally, this could cost up to £15.9 million (assuming a 60% medication price reduction).

**Conclusions:** There is evidence of variation in tolvaptan prescribing in the UK. A substantial proportion of patients eligible for tolvaptan were not initiated at the cohort-level, with evidence of variation between centres suggesting differences in treatment decision-making. A substantial proportion of patients initiated on tolvaptan were not eligible at the cohort-level, but there was limited evidence of variation between centres. Together, these findings raise questions regarding the potential consistency of clinical decision-making, equitable access to a sole disease-modifying therapy in a rare disease, alignment with national guidance, and effective use of healthcare resources.

## Introduction

Autosomal Dominant Polycystic Kidney Disease (ADPKD) is the most common inherited kidney disorder, affecting an estimated 12.5 million worldwide^1^ including 30,000-70,000 people in the United Kingdom (UK)^2^. It is characterised by progressive kidney cyst growth^3^, with approximately half of affected individuals reaching kidney failure by age 60^4^. ADPKD also impairs quality of life through abdominal pain and distension, and imposes substantial healthcare costs, primarily related to Kidney Replacement Therapy^5^.

Tolvaptan is the first disease-modifying therapy available for ADPKD and acts by antagonising vasopressin V2 receptors which reduces cyst fluid secretion and alters cystogenesis pathways. This mechanism explains the observed increase in urine output, thirst and nocturia which may impact quality of life. In pivotal clinical trials, including TEMPO 3:4 and REPRISE, tolvaptan treatment was associated with slower kidney enlargement and reduced decline in eGFR compared with placebo. However, trials also identified hepatotoxicity as a key safety concern requiring regular monitoring^6,7^.

Tolvaptan has been approved for adults with ADPKD who have evidence of rapidly progressing disease in over 43 countries worldwide^8^. Several international and national guidance documents have been published to support clinical decision-making and patient selection. These include the 2016 European Renal Association-European Dialysis Transplant Association, as well as expert consensus statements from countries such as Canada, Australia, New Zealand and Japan. Although specific criteria vary, these guidelines consistently emphasise identification of rapid disease progression using criteria such as historical eGFR decline, evidence of kidney growth on imaging, genetic risk and family history.

In the UK, the National Institute for Health and Care Excellence (NICE) approved tolvaptan in 2015 for ADPKD patients in CKD stages 2 or 3 with “rapidly progressive disease”^9^. The Renal Association issued recommendations for kidney centres providing care to people with ADPKD, defining rapid progression as an eGFR slope of >2.5ml/1.73m^2^/year over 5 years or >5ml/min/1.73m^2^ over 1 year, or total kidney volume increase of >5% per year. Additional measures for risk prediction may also be used based on imaging and genomic information^10^. Despite these recommendations, National Health Service (NHS) Digital data shows variation in tolvaptan reimbursement claims between NHS trusts^11^.

Such variation could reflect two potential departures from guidelines: patients eligible for tolvaptan who were not initiated and patients initiated on tolvaptan who did not meet eligibility criteria, which may be influenced by centre-level differences in prescribing. Variation may also arise from patient-level factors, such as case-mix or patient preferences, but for these to account for differences between centres they would need to vary systematically by centre and independently of other covariates. High-quality standardised performance measures adjust for factors outside the centre’s control, so that residual differences primarily reflect modifiable aspects of practice^12^.

Differences in centre-level prescribing may influence both access for eligible patients and exposure of ineligible patients to tolvaptan. Based on these considerations, we hypothesised a theoretical trade-off whereby kidney centres with high initiation rates may improve access but risk initiating patients not meeting eligibility criteria, while kidney centres with low initiation rates may limit inappropriate use but risk not initiating patients meeting eligibility criteria^13^.

This study aims to quantify variation in tolvaptan prescribing across UK kidney centres. Observed patterns may suggest potential unequal access to or treatment not aligned with guidelines for the sole disease-modifying therapy in a rare disease, with possible consequences for quality of life and healthcare costs.

## Materials and Methods

### Study Design and Population

A retrospective cohort study was conducted using RaDaR (National Registry of Rare Kidney Diseases) data to evaluate tolvaptan prescribing in adults with ADPKD across UK kidney centres. The cohort included adults ≥18 years with a recorded ADPKD diagnosis, eGFR results, and medication history via PatientView, which provides live linkage of patient-level clinical and laboratory data^14^. Participation is voluntary and medication history is added manually, meaning PatientView coverage varies by kidney centre. The study period (January 2016–September 2023) began three months following NICE approval, after which tolvaptan should be available at all NHS trusts^10^. Patients enrolled in tolvaptan clinical trials were excluded. Assessment of eligibility required sufficient laboratory data to derive eGFR slopes and medication history. Reporting followed the RECORD (Reporting of Studies Conducted using Observational Routinely-Collected Health Data) statement, an extension of the STROBE (Strengthening the Reporting of Observational Studies in Epidemiology) statement for observational studies using routinely collected data (Supplementary table 1)^15^. Full inclusion and exclusion criteria are detailed in Appendix 1. Data cleaning included range, consistency, and plausibility checks prior to analysis.

### Variables

Relevant patient characteristics in RaDaR include age, sex, ethnicity, CKD stage and eGFR. Medication characteristics for the year of eligibility and initiation were calculated as part of the study. Eligibility for tolvaptan was determined using Renal Association criteria recorded in the RaDaR database that was in place at the start of the study period: CKD stage 2 or 3 with an eGFR decline >2.5 ml/min/1.73m^2^/year over 5 years or >5 ml/min/1.73m^2^ over 1 year^16^. Individual eGFR slopes were calculated for each patient with a minimum of three eGFR measurements over at least 1 year required to derive a slope. EGFR measurements that were inconsistent with the patient’s overall longitudinal trajectory, such as abrupt isolated declines likely related to acute illness or measurement error, were excluded. Imaging, genotype and risk stratification based criteria were excluded due to inconsistent availability in the RaDaR database and relatively limited use in clinical practice according to expert consensus. Initiation was defined as the first recorded tolvaptan prescription within the study period. Key covariates included age, sex, CKD stage and year of eligibility or year tolvaptan was initiated, a priori as potential predictors of treatment being initiated and adherence to eligibility criteria.

## Statistical Analysis

### Descriptive Statistics

Continuous variables were summarised as mean (SD) or median (IQR), and categorical variables as counts and percentages. Distribution of patient characteristics across kidney centres was described.

### Patient-level Predictors

Univariable logistic regression was performed for each variable to assess its association with outcomes. Variables significant at p<0.05 or deemed clinically important were included in multivariable mixed-effects logistic regression models with kidney centre as a random intercept to account for clustering. Two logistic regression models were constructed:

- Initiation among eligible patients based on recorded eligibility criteria in RaDaR (binary outcome: 1 = initiated, 0 = not initiated)
- Eligibility based on recorded eligibility criteria in RaDaR among initiated patients (binary outcome: 1 = eligible, 0 = ineligible)

Odds ratios with 95% confidence intervals were reported for all variables.

### Kidney centre-level variation

Variation between centres was assessed using indirectly standardised ratios (ISRs). For each kidney centre, observed events (initiations or eligible patients) were compared to expected events based on patient-level characteristics, producing kidney centre-specific ratios. An ISR>100 indicates more events than expected for a kidney centre, whereas an ISR<100 indicates fewer events than expected. Funnel plots were used to visualise variation, with control limits at ±2 and ±3 standard deviations, corresponding to 95% and 99.8% confidence intervals respectively. Adjustment for overdispersion was applied to account for extra-binomial variation^17^.

### Health and cost implications

A deterministic budget impact and cost-effectiveness model was developed to estimate the consequences of patients eligible for tolvaptan based on recorded eligibility criteria recorded in RaDaR who were not initiated and patients initiated on tolvaptan who were not eligible based on recorded eligibility criteria recoded in RaDaR. This was conducted in accordance with the Principles of Good Practice for Budget Impact Analysis Report by the International Society for Pharmacoeconomics and Outcomes Research (ISPOR)^18^. Budget impact calculations incorporated estimates from the RaDaR analysis, published tolvaptan uptake rates and median treatment duration derived from two comparable United States cohorts in the absence of UK data (Appendix 2)^19,20^. To account for patient preferences, an uptake of 30.5% was assumed based on the findings of Calvaruso et al (2023). Five medication pricing scenarios were applied based on tolvaptan’s list price^9^ and hypothetical discounts that may be available through the patient access scheme (23%^21^, 60%^22^, 70%^23^ and 90%^23^).

A cohort-based simulation projected annual eGFR decline over 10 years using baseline eGFR from the study with published estimates of mean and standard deviation of annual eGFR decline in ADPKD patients^24^. Treatment effects were incorporated by adjusting the mean annual decline. For each year, the probability of reaching kidney failure was calculated using a normal approximation of eGFR decline and Z-scores relative to the kidney failure threshold. Expected eGFR trajectories for treated and untreated cohorts were calculated and cumulative kidney failure-free years were estimated accounting for pre-dialysis death. Gross potential savings from delaying progression to kidney failure were calculated using annual dialysis costs, adjusted for medication, monitoring costs and early discontinuation. Exact 95% confidence intervals for binomial proportions were calculated using the Clopper-Pearson method^25^. Results were expressed as the equivalent number of total hip replacements to aid interpretation.

### Statistical Software

All statistical analyses, including descriptive statistics and regression modelling, were conducted in Statistical Analysis System (SAS Institute, 2016, Version 9.4). Funnel and forest plots were generated using Microsoft Excel for Mac (Microsoft Corporation, 2019, Version 16.78.3). Deterministic budget impact and cost-effectiveness modelling was undertaken in the same software.

### Ethics

Ethics approval was granted by the Dulwich Research Ethics Committee in March 2024 (REC reference 24/PR/0106). Data from RaDaR were accessed for research purposes on 19/02/2024. DP is employed by the UK Renal Registry, the data controller for RaDaR. DP had access to the pseudonymised patient-level dataset. All other authors had access only to aggregated summary data and did not have access to identifiable individual participant information at any stage of the study.

## Results

### Cohort characteristics

A total of 8,135 patients were in the RaDaR ADPKD cohort from 89 kidney centres. Sequential application of inclusion criteria yielded 3,609 patients with complete laboratory and medication data for analysis from 72 kidney centres (Figure 1). Of the 4,526 excluded patients, the main reasons for exclusion were the absence of live linkage (32%, 1,457/4,526) and the absence of a recorded medication history (49%, 2,225/4,526). Participant exclusions were not evenly distributed but were driven by a small number of high-volume kidney centres accounting for a large proportion of missing data at each stage (see Appendix 3); 17 kidney centres did not have any medication history or live linkage data.

**Figure 1.**
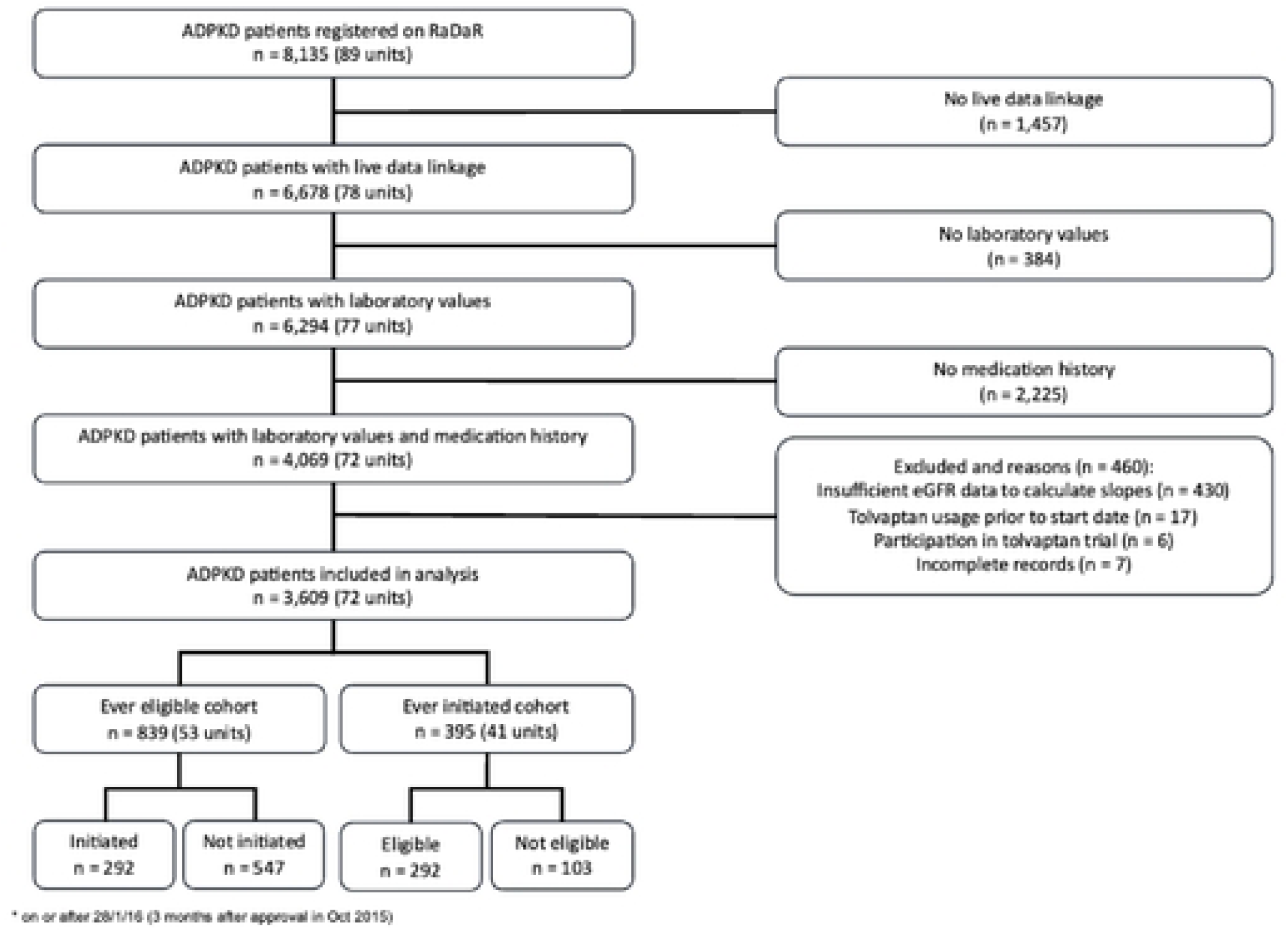
Participant flow diagram The mean number of patients per kidney centre was 50 (range 1 to 322). 51% female and 78% white ethnicity (Table 1).

**Table 1.**
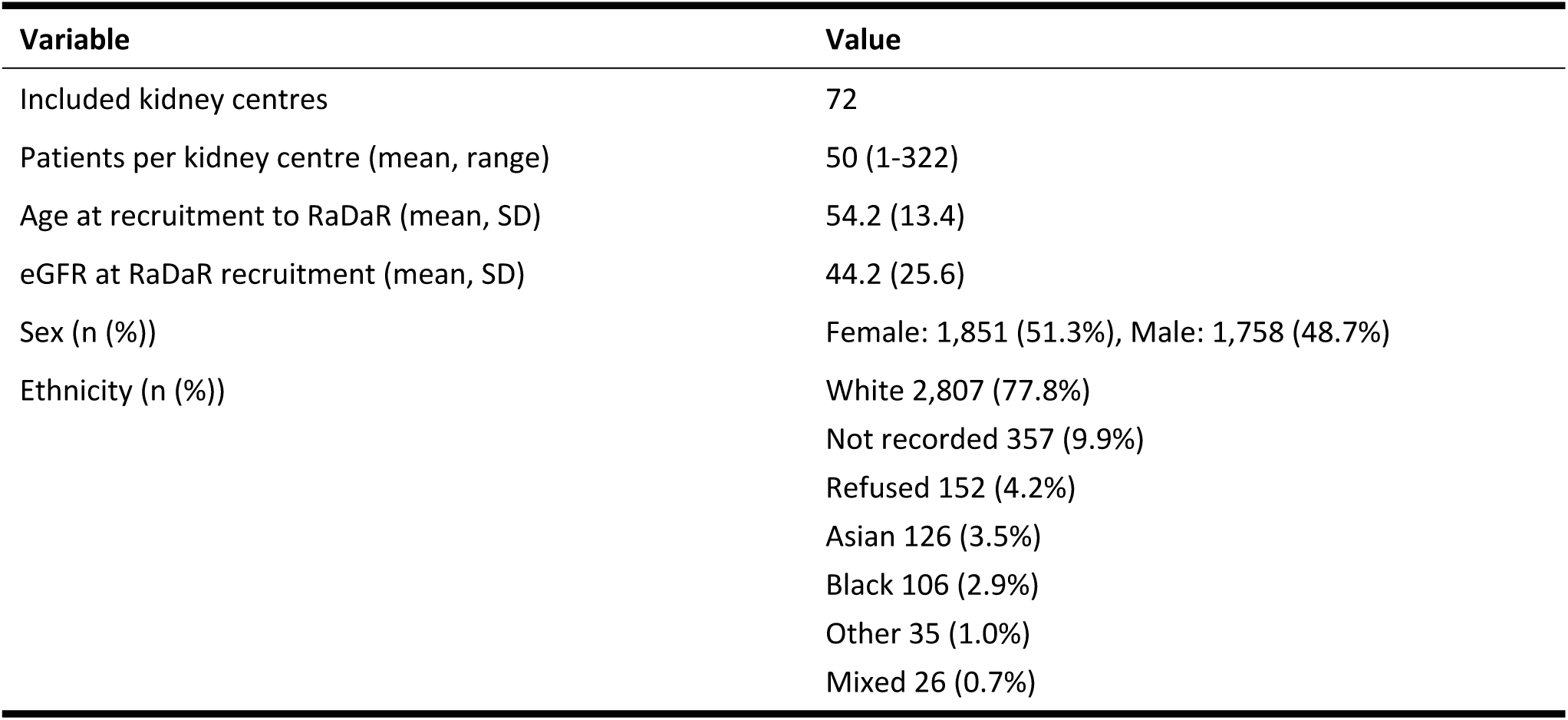
Characteristics of kidney centres and participants.

| Variable | Value |
| --- | --- |
| Included kidney centres | 72 |
| Patients per kidney centre (mean, range) | 50 (1-322) |
| Age at recruitment to RaDaR (mean, SD) | 54.2 (13.4) |
| eGFR at RaDaR recruitment (mean, SD) | 44.2 (25.6) |
| Sex (n (%)) | Female: 1,851 (51.3%), Male: 1,758 (48.7%) |
| Ethnicity (n (%)) | White 2,807 (77.8%)<br>Not recorded 357 (9.9%)<br>Refused 152 (4.2%)<br>Asian 126 (3.5%)<br>Black 106 (2.9%)<br>Other 35 (1.0%)<br>Mixed 26 (0.7%) |

### Patients eligible for tolvaptan who were not initiated

#### Patients eligible for tolvaptan, by initiation status

839 patients were eligible for tolvaptan with 65.2% (547/839) not being initiated on tolvaptan during the study period. Patients who were not initiated despite meeting eligibility criteria had slightly slower disease progression, with a 1-year eGFR decline that was on average 0.2 ml/min/1.73m^2^ slower than in initiated patients (−7.07 vs −7.27 ml/min/1.73m^2^ respectively) and a 5-year decline that was on average 0.22 ml/min/1.73m^2^ faster (−4.48 vs −4.26 ml/min/1.73m^2^ respectively). See Table 2 for the overview of initiated vs non-initiated patients eligible for tolvaptan.

**Table 2.**
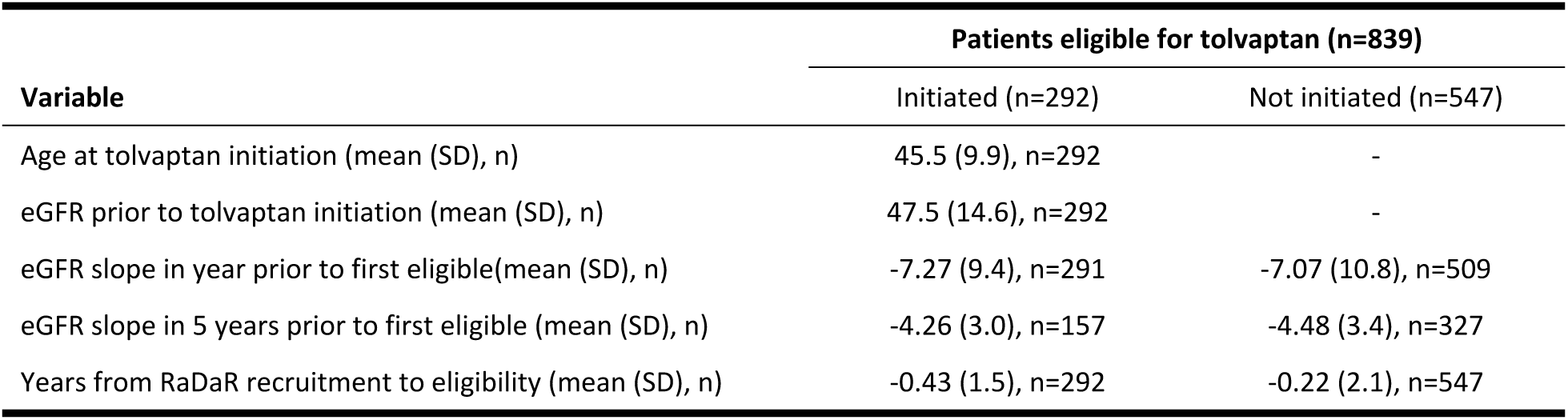
Characteristics of patients eligible for tolvaptan, stratified by initiation status.

| Variable | Patients eligible for tolvaptan (n=839) |  |
| --- | --- | --- |
|  | Initiated (n=292) | Not initiated (n=547) |
| Age at tolvaptan initiation (mean (SD), n) | 45.5 (9.9), n=292 | - |
| eGFR prior to tolvaptan initiation (mean (SD), n) | 47.5 (14.6), n=292 | - |
| eGFR slope in year prior to first eligible(mean (SD), n) | -7.27 (9.4), n=291 | -7.07 (10.8), n=509 |
| eGFR slope in 5 years prior to first eligible (mean (SD), n) | -4.26 (3.0), n=157 | -4.48 (3.4), n=327 |
| Years from RaDaR recruitment to eligibility (mean (SD), n) | -0.43 (1.5), n=292 | -0.22 (2.1), n=547 |

#### Patient-level predictors of initiation in eligible patients

In multivariable analysis, several factors were associated with the likelihood of initiation. Older age was associated with a lower likelihood of initiation (adjusted OR 0.964, 95% CI: 0.950-0.979, P<0.0001). Compared with CKD stages 1-2, individuals with CKD stages 3a and 3b had a higher likelihood of initiation (adjusted OR 3.77; 95% CI: 2.43-5.85 and adjusted OR 4.45; 95% CI: 2.83-7.02, respectively). Finally, the likelihood of being initiated varied with year of eligibility (global p=0.03). The highest likelihood of initiation was observed in 2017, it remained stable between 2018 and 2020, and then declined from 2021 onwards. Univariable and multivariable logistic regression results are summarised in Figure 2.

**Figure 2.**
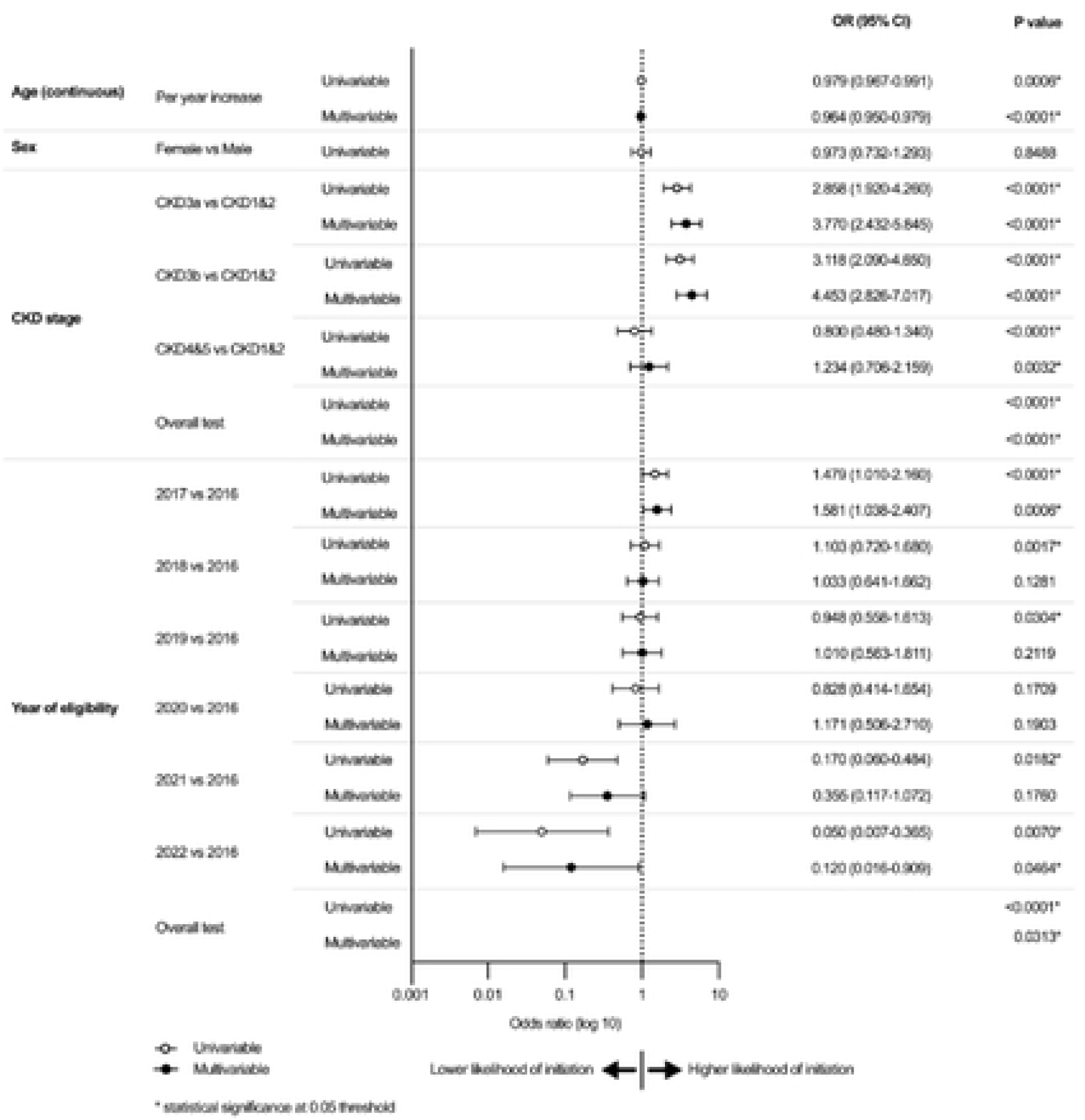
Forest plot of predictors for initiation in eligible patients

#### Initiation in eligible patients by kidney centres

Analysis of indirectly standardised ratios (ISRs) of tolvaptan initiation in eligible patients showed variation across the 53 included kidney centres (adjusted for age, CKD stage and year of eligibility). Only one kidney centre (1.9%) had significantly fewer patients initiated than expected among eligible patients, falling below the 3SD control limit. Five kidney centres (9.4%) were found to have significantly more patients initiated than expected among eligible patients, above the 2SD control limits. Adjusted funnel plots are included in Figure 3 and unadjusted funnel plots in Appendix 4.

**Figure 3.**
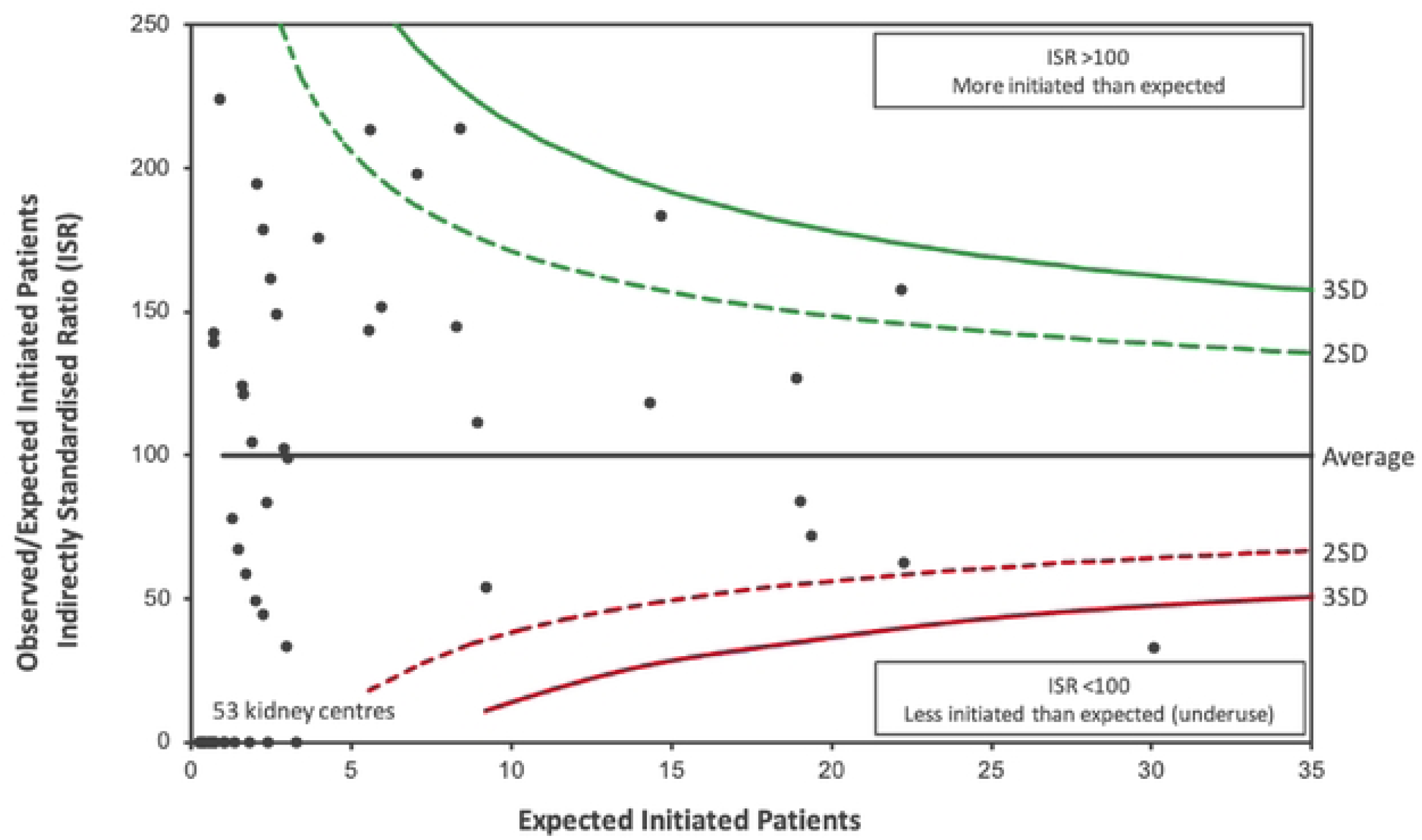
Tolvaptan initiation in-eligible patients by kidney centre; adjusted for age, CKD stage, and year of eligibility

### Patients initiated on tolvaptan who were not eligible

#### Patients initiated on tolvaptan, by eligibility

395 patients were initiated on tolvaptan with 26.1% (103/395) not eligible according to Renal Association criteria. Ineligible patients who were initiated had a higher mean age at initiation of 48.3 years compared with 45.5 years in eligible patients. Mean eGFR prior to initiation was lower in patients who were not eligible than those that were (47.5 vs 50.4 ml/min/1.73m^2^). Patients who were not eligible exhibited less rapidly progressive disease: the 1-year eGFR decline was 9.55 ml/min/1.73m^2^ slower than in patients who were not eligible than those that were (2.28 vs −7.27 ml/min/1.73m^2^) and the 5-year decline was 3.87 ml/min/1.73m^2^ slower (−0.39 vs −4.26 ml/min/1.73m^2^ per year). See Table 3 for the overview of not eligible vs eligible patients initiated on tolvaptan.

**Table 3.** Characteristics of patients initiated on tolvaptan, stratified by eligibility status.

| Variable | Patients initiated on tolvaptan (n=395) |  |
| --- | --- | --- |
|  | Eligible (n=292) | Not eligible (n=103) |
| Age at tolvaptan initiation (mean (SD), n) | 45.5 (9.91), n=292 | 48.3 (10.2), n=103 |
| eGFR prior to tolvaptan initiation (mean (SD), n) | 47.5 (14.6), n=292 | 50.4 (16.7), n=203 |
| eGFR slope in year prior to tolvaptan initiation (mean (SD), n) | -7.27 (9.4), n=291 | 2.28 (5.9), n=98 |
| eGFR slope in 5 years prior to tolvaptan initiation (mean (SD), n) | -4.26 (3.0), n=157 | -0.39 (2.0), n=94 |
| Years from RaDaR recruitment to eligibility (mean (SD), n) | -0.43 (1.5), n=292 | - |

#### Patient level predictors of eligibility in initiated patients

In univariable analysis, only CKD stage was a predictor of initiation in patients who were not eligible (overall test across CKD stages, p<0.0001). Compared to individuals with CKD stages 1 and 2, those with CKD3a were more likely to be eligible when initiated (OR 1.85, 95% CI 1.06-3.25, p=0.0006 and OR 3.23, 95% CI 1.84-5.66, p<0.0001, respectively), and CKD4 or CKD5 were less likely to be eligible (OR 0.12, 95% CI 0.04-0.37, p<0.0001), indicating greatest initiation in in late and then early stage CKD for people not eligible. Age, sex and year of initiation were not significantly associated. Multivariate adjustment for age, sex and year of initiation did not change the associations observed in univariable analyses; therefore, only univariable results are shown in Figure 4.

**Figure 4.**
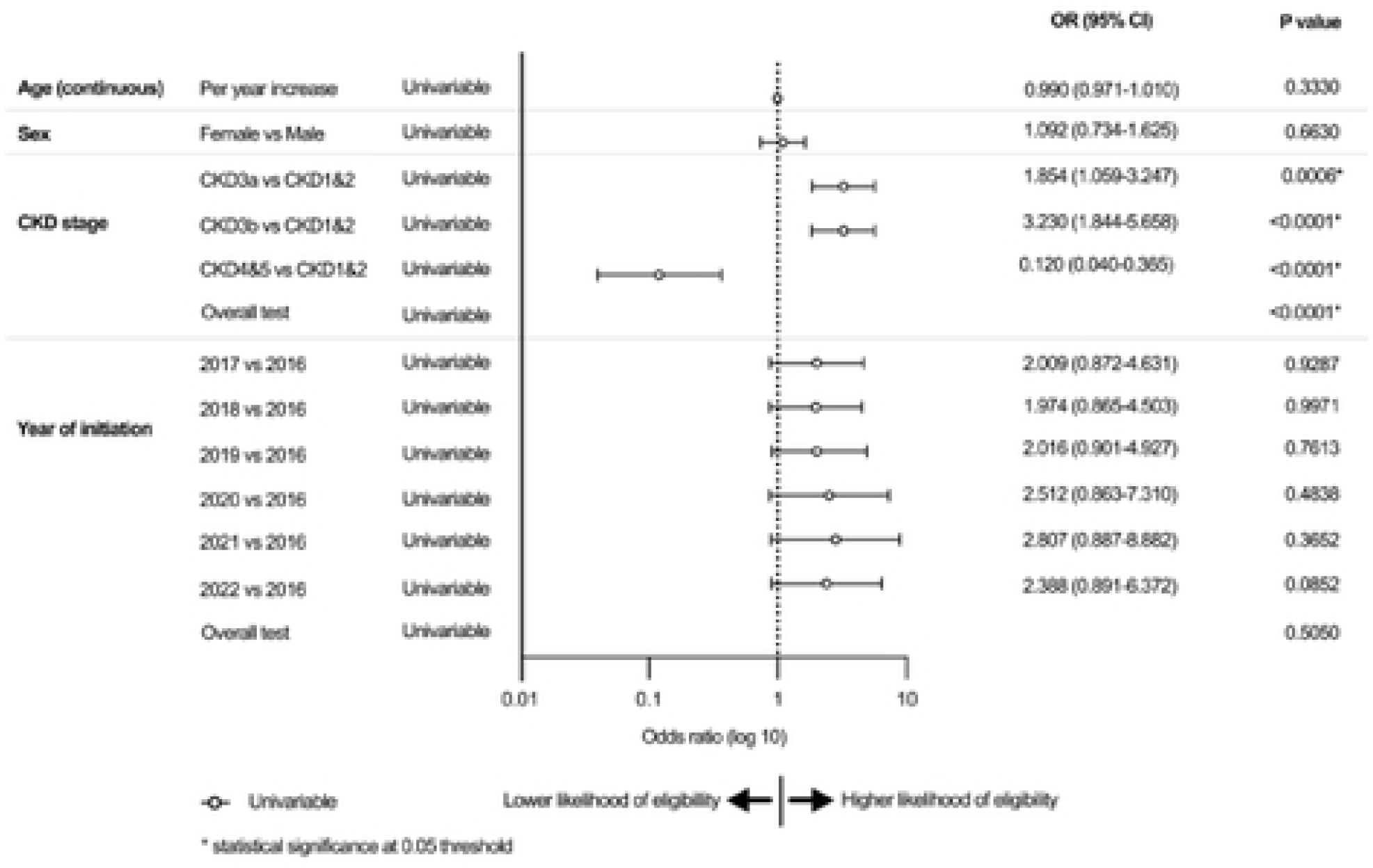
Forest plot of predictors of eligibility in initiated patients

#### Eligibility in initiated patients by kidney centres

Analysis of indirectly standardised ratios (ISRs) of tolvaptan eligibility in initiated patients across the 41 kidney centres included in the analysis (adjusted for CKD stage), revealed limited variation in practice. Only one kidney centre (2.4%) had significantly less eligible patients than expected among initiated patients, falling below the 2SD control limit. Adjusted funnel plots are shown in Figure 5 and unadjusted funnel plots in Appendix 5.

**Figure 5.**
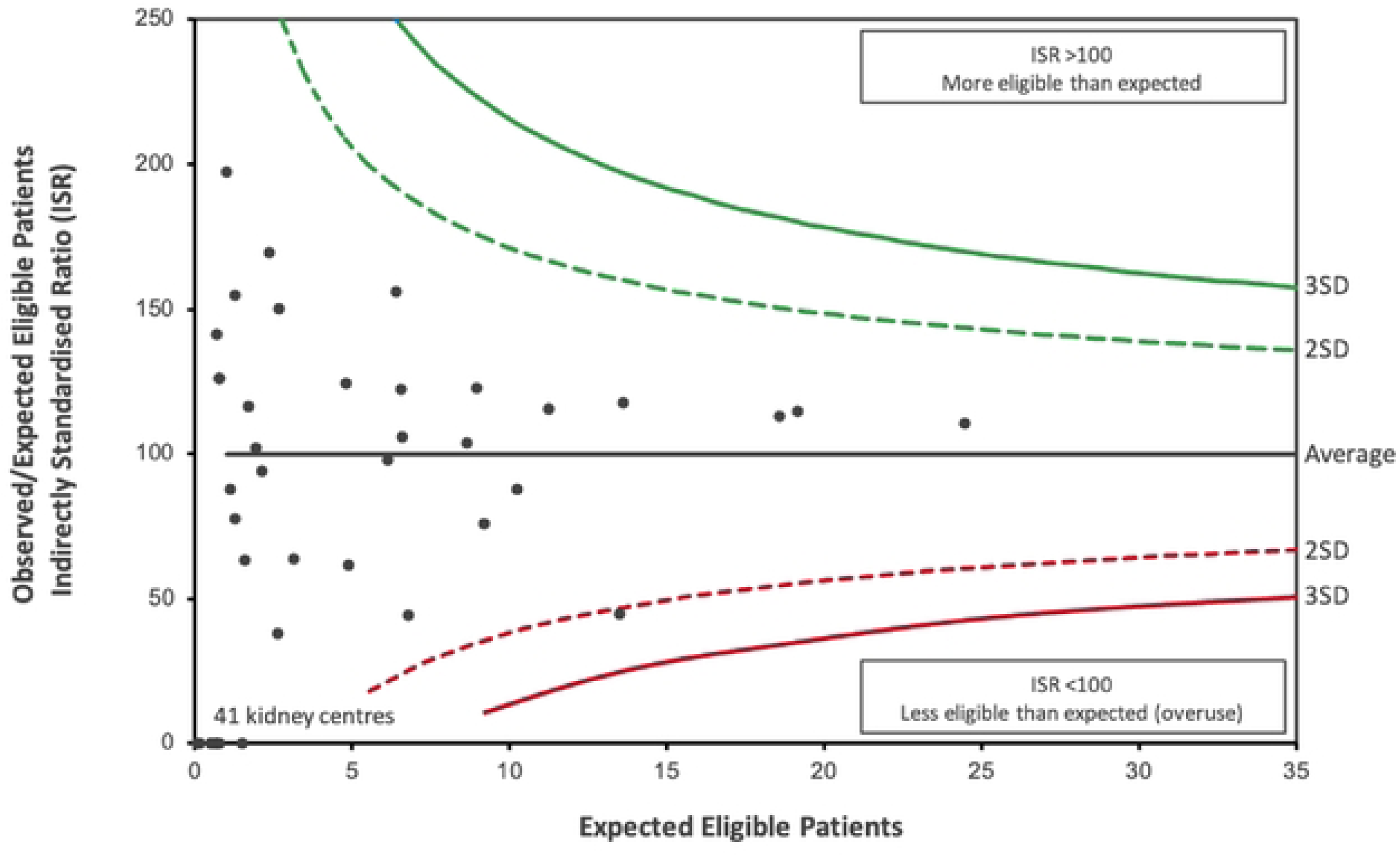
Tolvaptan eligibility in initiated patients by kidney centre; adjusted for CKD stage

#### Combined patterns of variation in tolvaptan prescribing

We hypothesised that there may be a theoretical trade-off whereby centres with high initiation rates may improve access for eligible patients but risk initiating a greater proportion of patients who were not eligible, while centres with low initiation rates may restrict initiation among patients who were not eligible but risk initiating a smaller proportion of patients who were eligible. However, the correlation between the proportion of eligible patients initiated and the proportion of initiated patients meeting eligibility criteria was weak and non-significant (r = 0.09, p = 0.63), indicating these behaviours occur independently (Appendix 6).

#### Health and cost implications of variation in practice

At a cohort-level, 65.2% of patients eligible based on recorded eligibility criteria for tolvaptan were not being initiated. Initiating treatment in this group could potentially save £78.2 million by delaying or avoiding dialysis assuming a patient uptake of 30.5% based on the study by Calvaruso *et al.* (2023), extrapolated to the UK ADPKD population. After accounting for medication and monitoring costs, discontinuation and pre-dialysis death, the associated net savings could range from £19.4 million at list price to £70.8 million with a 90% patient access scheme discount (Table 4). A 60% medication discount was applied, reflecting the typical level of patient access scheme agreements in the UK^22^ which would result in a real-world cost saving of £53.7 million. Considering the health impact of patients eligible for tolvaptan not starting treatment, the incremental quality-adjusted life years (QALYs) gained per patient treated with tolvaptan is estimated at 0.8977. Across the eligible ADPKD population not initiated on tolvaptan in the UK, this would equate to a total of 1,245 QALYs lost.

**Table 4.** Estimated net potential savings from initiating tolvaptan in eligible patients who were not initiated in the UK and estimated costs of initiating ineligible patients, by PAS discount level.

| PAS discount | Source | Starting eligible patients not previously initiated |  | Not starting ineligible patients previously initiated |  |
| --- | --- | --- | --- | --- | --- |
|  |  | Net potential savings (95% CI) (£ million) | Equivalent hip replacements (n) | Overall cost (95% CI) (£ million) | Equivalent hip replacements (n) |
| 0% discount | Company submission to NICE HTA<sup>26</sup> | 19.4 (15.1 – 24.2) | 2,084 | 38.1 (31.0 – 45.4) | 4,093 |
| 23% discount | Medicaid Drug Rebate program<sup>21</sup> | 32.5 (29.2 – 36.2) | 3,497 | 29.6 (24.1 – 35.3) | 3,178 |
| 60% discount | UK parliament document<sup>22</sup> | 53.7 (51.9 – 55.7) | 5,772 | 15.9 (12.9– 19.0) | 1,707 |
| 70% discount | Lower limit generic reduction<sup>23</sup> | 59.4 (58.0 – 60.9) | 6,387 | 12.2 (9.9– 14.5) | 1,310 |
| 90% discount | Upper limit generic reduction<sup>23</sup> | 70.8 (70.3 – 71.4) | 7,616 | 4.8 (3.9 – 5.7) | 515 |

At a cohort-level, 26.1% of patients initiated on tolvaptan were not eligible based on recorded eligibility criteria. Not initiating treatment in this group could potentially save between £4.8 million with a 90% discount and £38.1 million at list price if it was assumed prescriptions were not eligible according to all Renal Association eligibility criteria, extrapolated to the UK ADPKD population (Table 4). This takes into account medication and monitoring costs, assuming 2.74 years duration based on existing studies^19,20^. Based on anecdotal information regarding patient access schemes in the UK, the most likely real-world cost savings would be £15.9 million at a 60% discount. Appendices 7-11 and Supplementary tables 2-3 provide the underlying model and calculations supporting these results.

## Discussion

### Summary of key findings

In this national cohort of 3,609 patients with ADPKD across 72 UK kidney centres, tolvaptan prescribing in relation to nationally specified eligibility criteria exhibited systematic differences. Among 839 eligible patients, 65.2% (547/839) were not initiated. These patients demonstrated eGFR slopes comparable to those who were initiated. Kidney centre-specific rates of initiation in eligible patients also varied with one prescribing significantly less than expected to eligible patients and five prescribed significantly more after adjustment for case mix. Among 395 patients initiated on tolvaptan, 26.1% (103/395) were not eligible. These patients that were not eligible were older and had slower disease progression. Kidney centre-specific rates of eligibility in initiated patients showed minimal deviation, with only one kidney centre prescribing significantly less eligible patients than expected. We hypothesised a theoretical trade-off whereby kidney centres with high initiation rates may improve access but risk initiating patients not meeting eligibility criteria, while kidney centres with low initiation rates may limit inappropriate use but risk not initiating patients meeting eligibility criteria. However, the two behaviours appeared to occur independently.

Our findings could have important clinical and economic consequences. Patients eligible for tolvaptan who were not initiated may carry substantial opportunity cost: extrapolated UK-wide estimates suggest potential net savings of up to £53.7 million assuming a patient uptake of 30.5%, equivalent to 5,772 NHS total hip replacements, and could result in a loss of 1,245 QALYs. Conversely, patients initiated on tolvaptan who were not eligible could cost up to £15.9 million if it was assumed prescriptions were not eligible according to all Renal Association eligibility criteria, equivalent to 1,707 NHS total hip replacements. Overall, the largest health-system gains could potentially come from increasing initiation among eligible patients, as the potential gains would exceed the cost of treating a small number of patients were are not eligible.

### Comparison with Previous Studies

Previous national evaluations of tolvaptan use have relied on reimbursement or insurance claims aggregated by region^11,27^. While useful for broad trends, these datasets lack granularity to assess variation at the kidney centre level. Chong *et al.* (2022) reported a four-fold difference in prescribing across UK regions using NHS digital reimbursement data, but did not distinguish ADPKD from other indications for tolvaptan like heart failure or the syndrome of inappropriate antidiuretic hormone secretion^11^. Finally, in Japan, Inoue *et al.* (2020) reported substantial regional variation in tolvaptan prescribing with some prefectures prescribing up to 21 times more than others following its approval^28^. These international studies underscores that variation in tolvaptan prescribing is a broader phenomenon, not limited to the UK, and likely reflects differences in unmeasured factors. However, none of these studies have reported prescribing by nationally specified eligibility criteria or considered the economic and health consequences of variation observed in practice.

The 65.2% of patients eligible for tolvaptan who were not initiated on tolvaptan could be related to patient refusal which was not captured in RaDaR data and has not been published for the UK ADPKD population. In Canada, Calvaruso *et al.* (2023) reported a 30.5% uptake among high-risk patients at a single kidney centre using Mayo Imaging Classes which was used in our health and economic analysis^27^.. However, for such patient-level factors to fully explain observed centre-level variation they would have to vary systematically across centres and independently of the covariates included. While observed variation may therefore overestimate missed opportunities, residual differences after adjustment are still likely to reflect centre-level practice^12^. The 26.1% of patients initiated on tolvaptan who were not eligible could be related to true prescribing outside of established guidelines or based on the other eligibility criteria that were previously thought to be used infrequently in clinical practice.

An unexpected finding was a positive 1-year eGFR slope in patients initiated on tolvaptan who were not eligible compared with eligible patients (2.28 vs −7.27 ml/min/1.73m^2^). This may reflect patients who previously met eGFR slope criteria but no longer fulfilled them at the time of initiation. Alternatively, tolvaptan may have been initiated earlier in the disease course during a phase of hyperfiltration, as demonstrated by the Consortium for Radiologic Imaging Studies of Polycystic Kidney Disease (CRISP) study which showed that early stage ADPKD patients with TKV <750ml can have positive eGFR slopes^29^. In such cases, clinicians may be considering additional risk factors outside of the formal eligibility criteria such as family history, anticipated rate of progression or accommodating patient requests^30^.

Although RaDaR captures primarily patient-level data, it cannot fully explain the observed variation in our study. Previous evidence syntheses in both guideline implementation in rare diseases and prescribing decision-making in chronic diseases have reported that practices are influenced by factors at the patient, prescriber, healthcare-system and external organisations^30,31^. However, it is unclear to what extent and in what ways these factors contribute to the variation in prescribing patterns observed in our cohort.

### Strengths and Limitations

This is the first national-level analysis of real-world tolvaptan prescribing using a large ADPKD cohort, reporting patterns at both the patient and kidney centre-level. The inclusion of economic modelling is an additional strength, quantifying the potential health and cost implications associated with prescribing patterns. This provides policy-relevant insights into the resource implications of variation. Identification of kidney centres with differing prescribing patterns may also help guide targeted qualitative research.

Three limitations should be noted. First, there was missing data but this was largely at the level of whole kidney centres rather than individual patients. Even among included centres, patient numbers were relatively small in some cases which could reduce the precision of centre-level estimates. Other authors have recognised similar limitations in registry analyses^32–34^. The Real-World Evidence Framework published by NICE recognised the value of using routinely collected clinical data to provide insights into treatment patterns, outcomes and resource use that can inform economic analyses, even when data is less complete or detailed than clinical trial data^35^. Second, patient-level reasons for non-initiation, such as preferences, reproductive considerations, or lifestyle impact from aquaretic adverse effects, were not captured. Third, although eligibility for tolvaptan was only based on eGFR slopes due to the lack of imaging and genetic data, this is in line with UK prescribing practice.

### Implications and Recommendations

While these findings are derived from UK data, the underlying issues of high-cost therapy access, guideline adherence and variation in prescribing are relevant to ADPKD care in other healthcare systems. Beyond ADPKD, this approach demonstrates how national registries can be used to characterise treatment variation and resource implications in rare diseases more broadly.

Our findings may have important implications for patient outcomes and NHS resource use. Patients eligible for tolvaptan who were not initiated may reflect patient refusal or missed opportunities to access a sole disease-modifying therapy, while patients initiated on tolvaptan when not eligible suggest that prescribing may sometimes occur outside published recommendations or that other eligibility criteria thought to be rare used are being applied in clinical practice.

Further research using qualitative methods is needed to better understand the clinical, organisational and contextual factors driving the observed variation in tolvaptan prescribing, and explore the consequences of diverging from prescribing guidance.

## Conclusions

There is evidence of variation in tolvaptan prescribing in the UK. A substantial proportion of patients eligible for tolvaptan were not initiated at the cohort-level, with evidence of variation between centres suggesting differences in treatment decision-making. A substantial proportion of patients initiated on tolvaptan were not eligible at the cohort-level, but there was limited evidence of variation between centres. Together, these findings raise questions regarding the potential consistency of clinical decision-making, equitable access to a sole disease-modifying therapy in a rare disease, alignment with national guidance, and effective use of healthcare resources.

## Data Availability

The data used in this study are available from the RaDaR registry but are not publicly available due to governance and ethical restrictions. Access to the data requires approval through the RaDaR data access process and appropriate institutional permissions.

## Acknowledgements

We gratefully acknowledge the UK Renal Registry and RaDaR (Rare and Genetic Kidney Disease Registry) for access to the data used in this study and the provision of statistical support. We also thank The Healthcare Improvement Studies (THIS) Institute for their support and contribution to this work.

## Author contributions

MG conceptualisation, funding acquisition, methodology, formal analysis, visualisation, writing – original draft preparation, writing – review & editing

DP data curation, formal analysis, resources, software, visualisation

AOC supervision, writing – review & editing

ACO conceptualisation, supervision, writing – review & editing

JF conceptualisation, funding acquisition, methodology, supervision, writing – review & editing

